# Machine learning to identify socio-behavioural predictors of HIV positivity in East and Southern Africa

**DOI:** 10.1101/2020.01.27.20018242

**Authors:** Erol Orel, Rachel Esra, Janne Estill, Stéphane Marchand-Maillet, Aziza Merzouki, Olivia Keiser

## Abstract

**Background:** There is a need for high yield HIV testing strategies to reach epidemic control. We aimed to predict the HIV status of individuals based on socio-behavioural characteristics.

**Methods:** We analysed over 3,200 variables from the most recent Demographic Health Survey from 10 countries in East and Southern Africa. We trained four machine-learning algorithms and selected the best based on the f1 score. Training and validation were done on 80% of the data. The model was tested on the remaining 20% and on a left-out country which was rotated around. The best algorithm was retrained on the variables which were most predictive. We studied two scenarios: one aiming to identify 95% of people living with HIV (PLHIV) and one aiming to identify individuals with 95% or higher probability of being HIV positive.

**Findings:** Overall 55,151 males and 69,626 females were included. XGBoost performed best in predicting HIV with a mean f1 of 76·8% [95% confidence interval 76·0%-77·6%] for males and 78·8% [78·2%-79·4%] for females. Among the ten most predictive variables, nine were identical for both sexes: longitude, latitude and, altitude of place of residence, current age, age of most recent partner, total lifetime number of sexual partners, years lived in current place of residence, condom use during last intercourse and, wealth index. Model performance based on these variables decreased minimally. For the first scenario, 7 males and 5 females would need to be tested to identify one HIV positive person. For the second scenario, 4·2% of males and 6·2% of females would have been identified as high-risk population.

**Interpretation:** We were able to identify PLHIV and those at high risk of infection who may be offered pre-exposure prophylaxis and/or voluntary medical male circumcision. These findings can inform the implementation of HIV prevention and testing strategies.

**Funding:** Swiss National Science Foundation.

## Introduction

In order to reach epidemic control by 2030, the Joint United Nations Programme (UNAIDS) have set fast track targets to rapidly scale up effective HIV services.^1^ One of the aims is to ensure that 95% of the approximately 38 million people living with HIV (PLHIV) are aware of their HIV status and that 95% of those with HIV positive diagnoses are on treatment.^2^

People in East and Southern Africa are disproportionately burdened by HIV, constituting more than half of the global PLHIV with 20·6 million people currently estimated to be HIV positive.^2^ As of 2018, 85% of PLHIV in this region were aware of their HIV status, of whom 79% were accessing treatment.^3^ In addition, 25% of new HIV infections in East and Southern Africa were concentrated among key populations such as female sex workers, men having sex with men, prisoners and, people who inject drugs.^3^

HIV is transmitted within a complex network that is influenced by biological, behavioural and, social factors. In East and Southern Africa, there is large geographical variation in the distribution of the HIV epidemic.^4^ In order to identify populations at a high risk of infection, global HIV prevention efforts have shifted toward optimizing resource allocation by considering geographical data as a way of increasing program impact and efficiency.^5^

Machine learning algorithms have the power to substantially enhance HIV prevention and detection, increasing the prediction capability by processing large amounts of data of a different nature. This methodology has been implemented to establish patterns of HIV risk behaviour, to optimise HIV treatment modalities and, to identify high-risk individuals for targeted interventions from a number of novel data sources.^6–15^

As more PLHIV are diagnosed, finding persons with undiagnosed HIV becomes progressively more difficult and expensive. Hence, resource constraints and potential funding shortages have resulted in demands for differentiated high yield testing strategies in parallel to provider-initiated HIV testing and counselling (PITC).^14,16,17^ We therefore aimed to compare different machine learning algorithms to identify new key populations based on a variety of socio-behavioural characteristics. These insights intend to both inform targeted case-finding strategies as well as identify high risk HIV negative individuals eligible for prevention services such as voluntary medical male circumcision (VMMC) and/or pre-exposure prophylaxis (PrEP).

## Methods

### Data

Since 1984, the Demographic and Health Surveys (DHS) program has provided technical assistance for over 400 surveys in more than 90 countries, advancing global understanding of health and population trends in developing countries.^18^ DHS are nationally-representative household surveys that provide data for a wide range of monitoring and impact evaluation indicators on health and nutrition. Standard DHS surveys have large sample sizes (usually between 5,000 and 30,000 households) and are typically conducted every five years.^19^ We used the most recent DHS surveys at or after 2013 of ten East and Southern African countries (Table A1) with a generalised HIV epidemic: Angola, Burundi, Ethiopia, Lesotho, Malawi, Mozambique, Namibia, Rwanda, Zambia and, Zimbabwe. Male and female’s datasets were combined separately with geographic position of groups of households where the individuals live and HIV test results. We then merged the ten countries and obtained two datasets containing 68,979 males and 83,910 females with 527 and 3,213 variables, respectively.

### Data pre-processing, model validation and, algorithm selection

We compared four machine learning algorithms for the prediction of the HIV status of an individual; a penalized logistic regression (Elastic Net),^20^ a generalized additive model (GAM), ^21^ a support vector machine (SVM) and,^22^ a gradient boosting tree (XGBoost).^23^ The Elastic Net and the GAM are among the most widely used classification methods in biology and medicine, SVM is a very common machine learning algorithm and XGBoost is a decision-tree based ensemble which has gained a lot of attraction since its development few years ago.

The first part of the analyses was done in several steps for each of the four algorithms, and separately for males and females (Figure 1). In the data pre-processing step (Figure 1, step 1), we first cleaned and transformed data from the ten countries into numerical values (Table A2). Only persons for whom the HIV status was either positive or negative were included in the analysis. The cleaned datasets included 55,151 males and 69,626 females with 84 and 122 variables, respectively; 73 variables were common for both sexes (Table A3). Since we wanted to test the generalizability of our model, one country was left out for later testing, and the left-out country was rotated around. We then split each of the data from nine countries combined in an 80% training sample and a 20% test sample. Missing values were imputed using multiple imputation by chained equations (MICE) (as detailed in appendix) and data were further harmonized and scaled.^24^

**Figure 1:**
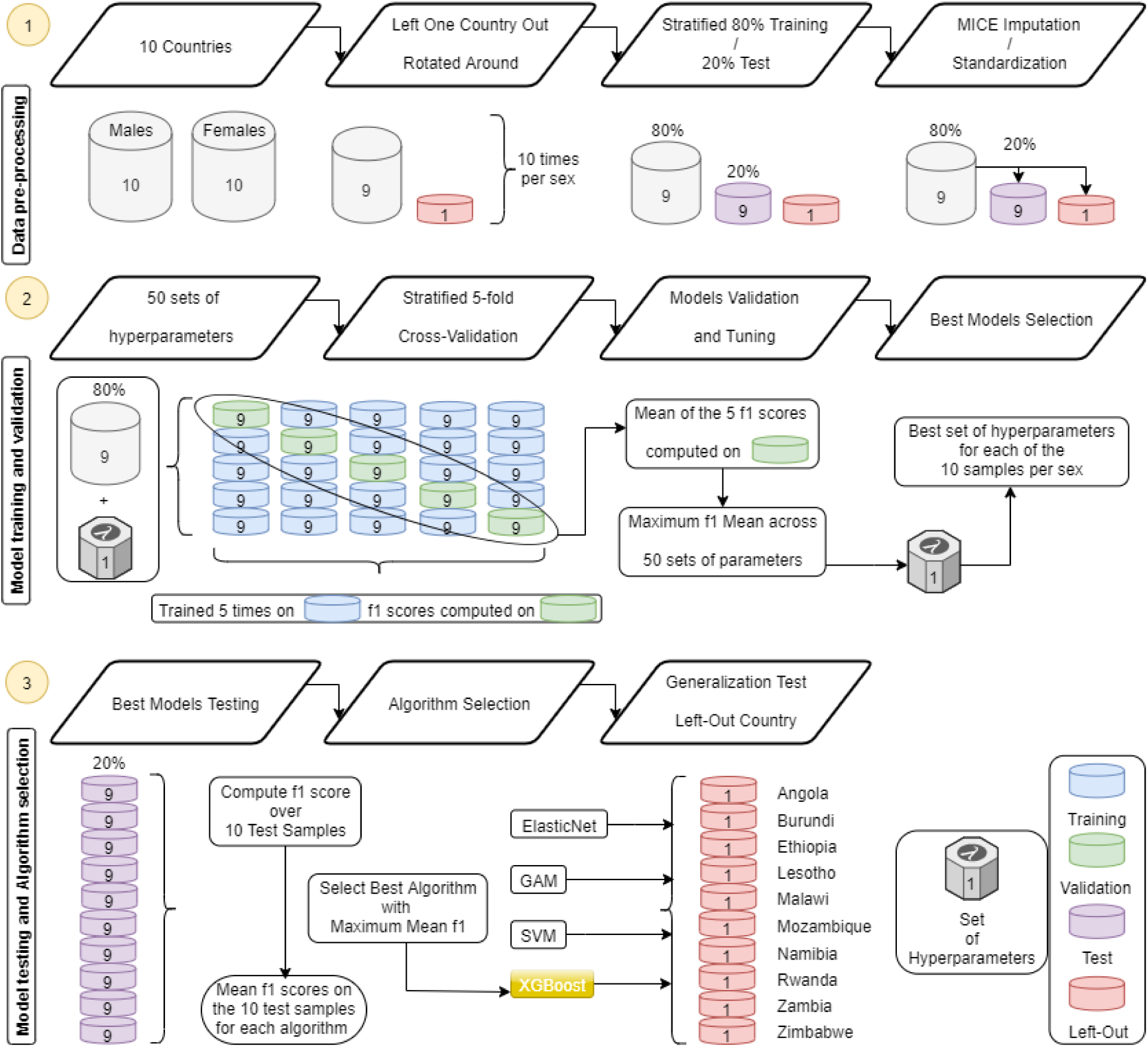
Diagram explaining the first part of the analyses. All steps are detailed in the method section.

Training and validation were done using five-fold cross-validation on 50 sets of hyperparameters randomly chosen from a grid (Figure 1, step 2). For each of these 50 sets, we calculated the mean f1 scores across the five validation sub-samples and selected the set of hyperparameters that gave the highest value. The f1 score combines the sensitivity and the PPV in a harmonic mean.^25^ Our primary interest was to find a large number of HIV positive persons (sensitivity or recall) with a high yield (precision or positive predictive value (PPV)). The probability threshold to classify if someone is considered HIV positive was set at 50%.

Once the best models of each algorithm were obtained, we calculated the f1 scores on the ten 20% test and left-out country samples which were not used to train and validate the model (Figure 1, step 3). We averaged these f1 scores in order to compare the different algorithms and the maximum mean f1 on the 20% test samples allowed us to select the best one.

### Variables selection, direction of association and, calibration of two scenarios

For the second part of the analysis, where no country was left out, only the selected algorithm was trained and validated again using a random search over 250 sets of parameters (instead of 50) with the same five-fold cross-validation scheme than previously. The first estimation was performed using all variables. We compared the f1 score, the sensitivity and, the PPV using MICE imputation (models M1 and F1 for males and females, respectively) with a different imputation method within the algorithm,^23^ that considerably simplified the engineering process (models M2 and F2).

We then performed a sequential forward floating selection (SFFS) using the best imputation method on the 80% training samples and calculated the f1 scores for different number of variables. We selected the subset of variables for which the f1 scores plateaued and assessed the direction of the association between these variables and the probability of being HIV-positive using Shapley values.^26^

We retrained the best algorithm with the above defined subsets of variables (models M3 and F3) and also on a minimal subset common for both sexes (models M4 and F4). The f1 scores, the sensitivity and, the PPV were compared to the ones obtained for M1, M2, F1 and, F2. We computed the Precision-Recall (PR) and the Threshold-Scores curves (TS) for our preferred model per sex. The PR curve displays the PPV for different sensitivities. This curve is not influenced by imbalanced datasets and is therefore preferred over ROC curve.^27^ The TS curve, highlights the PPV, the sensitivity and, the f1 score for varying thresholds of classifying if someone is HIV positive.

We then tested two scenarios: for the first scenario, the sensitivity was set to 95%, equivalent to 95% of PLHIV knowing their status. We selected the threshold that corresponds to this sensitivity and reported the corresponding precision and number of individuals to be tested. For the second scenario, we identified a population for which the predicted probability of being HIV positive was 95% or higher. We considered that these individuals are either HIV positive targets for high yield testing strategies or HIV negative individuals who would be ideal candidates for prevention services.

All analyses were performed in Python version 3.7.4. The code is available on https://gitlab.com/Triphon/predicting_hiv_status.

## Results

Overall, 55,151 males and 69,626 females were analysed with an HIV positivity ranging from 0·8% among males in Ethiopia to 33·3% among females in Lesotho. The overall HIV positivity was 8·0% (4,417 individuals) for males and 11·5% (8,011 individuals) for females. Persons aged 25 to 34 years represented the largest age group representing 35·9% of females and 31·9% of males. About two-thirds of people lived in rural areas (Table A4).

### Algorithms’ performance on the test samples

Figure 2 shows the performance of the four algorithms. XGBoost had the highest f1 scores on all 20 test samples (ten per sex) with a mean f1 score of 76·8% [95% confidence interval (CI) 76·0%-77·6%] for males and 78.8% [78·2%-79·4%] for females. For SVM, the mean f1 score for males was 69·2% [68·2%-70·2%] and 74·6% [73·7%-75·5%] for females and for Elastic Net, the mean f1 score was 32·6% [31·8%-33·4%] for males and 41·5% [40·3%-42·7%] for females. GAM performed worst with a mean f1 score of 26·2% [25·0%-27·4%] for males and 39·8% [38·1%-41·5%] for females (see Table A5i to Table A5iv).

**Figure 2:**
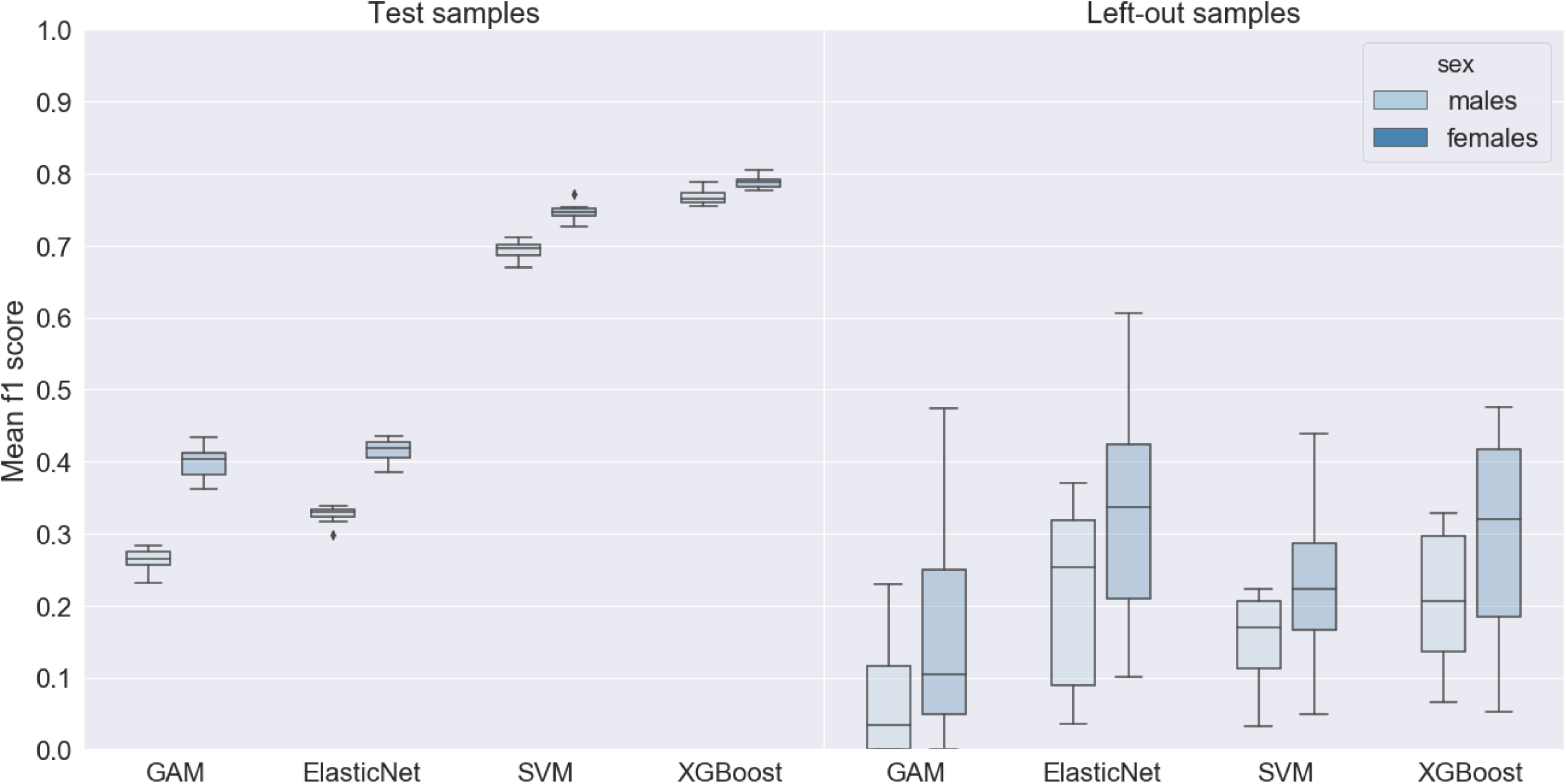
Boxplot of the f1 scores for the 4 algorithms on the test and left-out samples per sex. Generalized Additive Model (GAM), Support Vector Machine (SVM).

### Algorithms’ performance on the left-out country samples

The performance of the algorithms on the ten left-out samples was substantially lower than on the test samples and the f1 scores varied more widely (Figure 2). The mean f1 score was best for Elastic Net with 21·4% [12·3%-30·5%] for males and 32·6% [21·2%-44·0%] for females followed by XGBoost with 20·9% [14·3%-27·5%] and 29·8% [19·0%-40·6%], respectively. For SVM, the mean f1 score was 15·4% [10·9%-19·9%] for males and 22·3% [14·1%-30·5%] for females. Again, GAM performed worst with a mean f1 scores of 6·6% [0·9%-12·1%] and 17·1% [4·4%-29·8%] (see Table A5i to Table A5iv). The algorithms performed better in countries with higher HIV positivity.

### Best algorithm’s performance on the complete datasets

We selected the best performing algorithm, XGBoost, for the second part of the analysis, where no country was left out. The results on all variables using the two different imputation methods are shown in Table 1. For both sexes, the XGBoost imputation (M2 and F2) resulted in slightly higher f1 scores compared to the MICE imputation (M1 and F1). The f1 scores on the validation samples were 75·5% [73·7%-77·3%] vs 74·9% [73·3%-76·5%] for males and 76·1% [74·9%- 77·3%] vs 75·5% [74·6%-76·4%] for females. Given the above results and the simplicity of the XGBoost imputation, we used this imputation for further analyses (i.e. models M3, F3, M4 and, F4).

**Table 1:** Results per sex of the XGBoost algorithm for different imputation methods and sets of variables. True positive (TP), False negative (FN), False positive (FP), True negative (TN), Positive Predictive Value (PPV).

|  |  | TP | FN | FP | TN | f1 score | Sensitivity | PPV |
| --- | --- | --- | --- | --- | --- | --- | --- | --- |
| Complete with MICE imputation (Model M1) | Validation | | | | | 74.9% ($\pm 1.6\%$) | 71.2% ($\pm 2.9\%$) | 79.1% ($\pm 0.8\%$) |
|  | Test | 627 | 256 | 164 | 9,984 | 74.9% | 71.0% | 79.3% |
| Complete with MICE imputation (Model F1) | Validation | | | | | 75.5% ($\pm 0.9\%$) | 75.4% ($\pm 1.6\%$) | 75.6% ($\pm 0.5\%$) |
|  | Test | 1,264 | 338 | 375 | 11,949 | 78.0% | 78.9% | 77.1% |
| Complete with XGBoost imputation (Model M2) | Validation | | | | | 75.5% ($\pm 1.8\%$) | 69.6% ($\pm 2.2\%$) | 82.5% ($\pm 2.2\%$) |
|  | Test | 617 | 266 | 122 | 10,026 | 76.1% | 69.9% | 83.5% |
| Complete with XGBoost imputation (Model F2) | Validation | | | | | 76.1% ($\pm 1.2\%$) | 75.5% ($\pm 1.7\%$) | 76.8% ($\pm 1.2\%$) |
|  | Test | 1,279 | 323 | 379 | 11,945 | 78.5% | 79.8% | 77.1% |
| 15 variables with XGBoost imputation (Model M3) | Validation | | | | | 73.7% ($\pm 2.9\%$) | 67.9% ($\pm 2.5\%$) | 80.7% ($\pm 3.7\%$) |
|  | Test | 605 | 278 | 129 | 10,019 | 74.8% | 68.5% | 82.4% |
| 27 variables with XGBoost imputation (Model F3) | Validation | | | | | 75.6% ($\pm 1.2\%$) | 70.0% ($\pm 1.2\%$) | 82.2% ($\pm 1.7\%$) |
|  | Test | 1,212 | 390 | 234 | 12,090 | 79.5% | 75.7% | 83.8% |
| 9 variables with XGBoost imputation (Model M4) | Validation | | | | | 72.9% ($\pm 2.3\%$) | 65.6% ($\pm 1.6\%$) | 81.9% ($\pm 3.9\%$) |
|  | Test | 595 | 288 | 124 | 10,024 | 74.3% | 67.4% | 82.8% |
| 9 variables with XGBoost imputation (Model F4) | Validation | | | | | 72.4% ($\pm 1.2\%$) | 68.5% ($\pm 1.4\%$) | 76.8% ($\pm 1.6\%$) |
|  | Test | 1,184 | 418 | 249 | 12,075 | 78.0% | 73.9% | 82.6% |

Multiple Imputation by Chained Equations (MICE)
( $\pm \%$ ): 95% Confidence Interval

### Variables selection and direction of associations

Figure 3A and 3B show the result of the SFFS procedure which was used to select a subset of most relevant variables. The f1 scores plateaued above 99·6% with 15 variables for males and above 97·6% with 27 variables for females. Figure 3C and 3D show the 15 and 27 key variables of individual HIV status for males and females. Among these top ten most predictive variables, nine were identical: geographic position (longitude, latitude and, altitude), current age, age of most recent partner, total lifetime number of sexual partners, years lived in current place of residence, condom used during last sexual intercourse with most recent partner, and, a wealth index from the DHS which combines numerous wealth-related variables such as household assets and utility services.^28^ The age at first sexual intercourse ranked tenth for males and twentieth for females. The Rohrer’s index (an estimate of obesity) ranked sixth for females, but was not available for males. Among the fifteen most predictive variables, four were specific for either males or females (‘number of women fathered children with’ and ‘respondent circumcised’ for males and ‘currently breastfeeding’ and ‘fertility preference’ for females). Other females-specific characteristics included ‘time to get to water source’ and ‘entries in birth history’.

**Figure 3:**
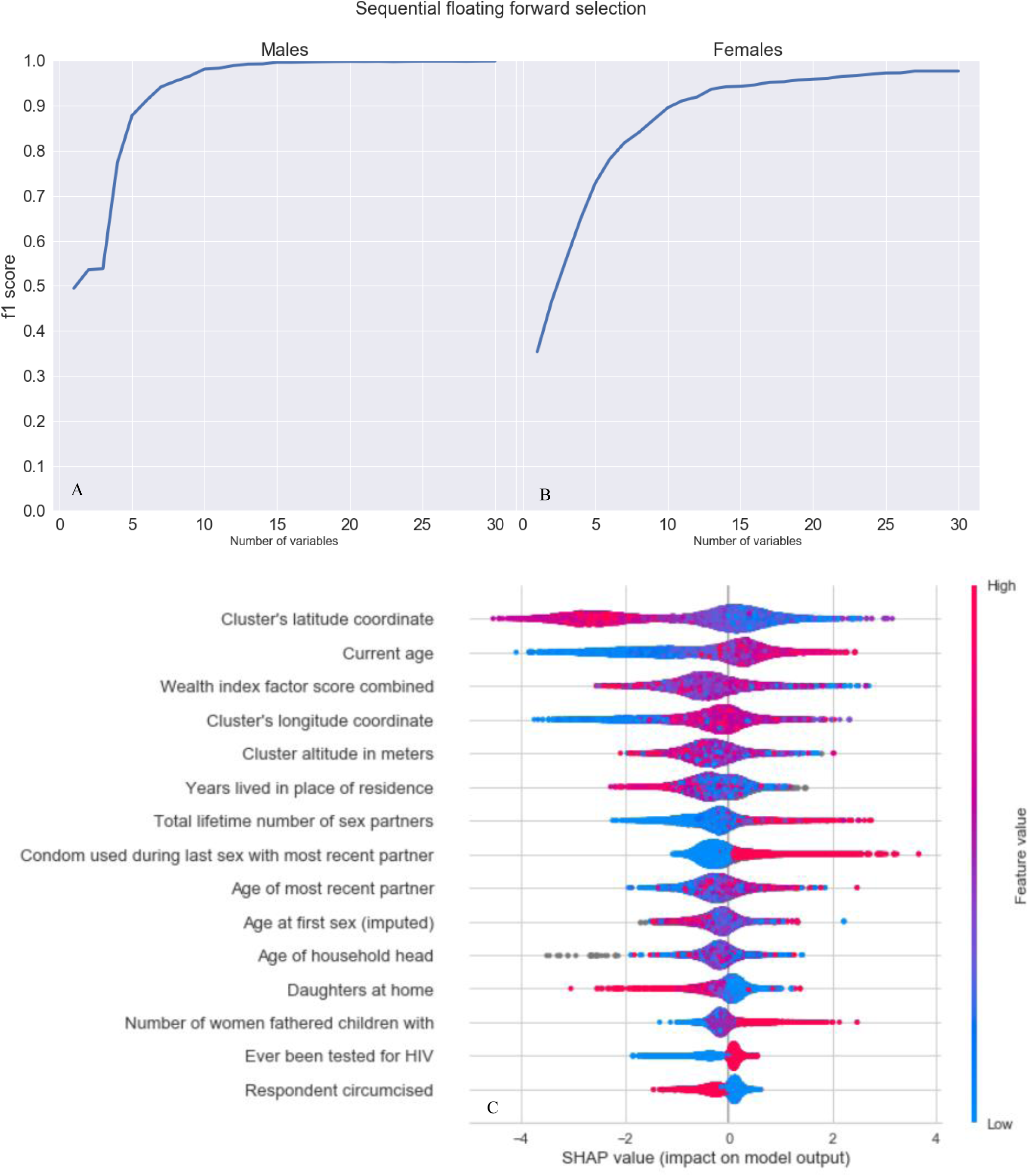
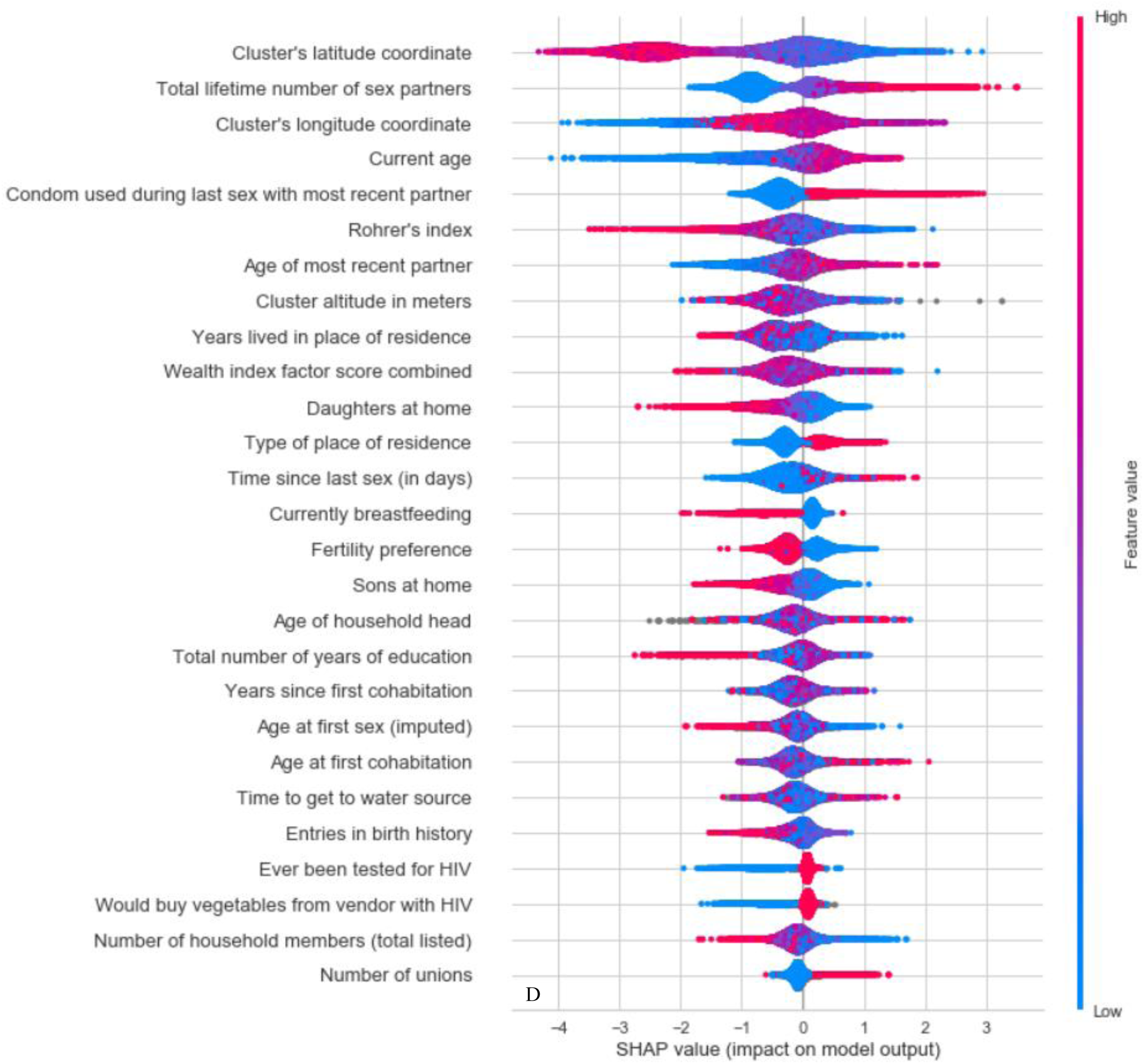
Sequential floating forward selection (SFFS) and Shapley values. The (SFFS) procedure was implemented (A + B) to determine the saturation point for variable selection base on the f1 score. This resulted in the selection of the 15 and 27 most important variables for males and females, respectively. The variables are displayed below (C + D) sorted by importance from top to bottom (from the highest Shapley value to the lowest). The blue and red colours represent the value range of the variable (e.g. blue represents low value range of the variable). For example, the older the age the more likely the persons will be HIV positive.

Figure 3A and 3B highlight the direction of the association between each variable and the probability of HIV positivity. Among the nine common predictive variables for both sexes, older age, older age of most recent partner, a higher number of total lifetime number of sexual partners, condom used during last sexual intercourse with most recent partner and, longitude were positively associated with the probability of HIV positivity for most individuals. A higher wealth index, a larger latitude coordinate of the residence, altitude and, more years lived in place of residence were mainly negatively associated with HIV positivity.

### Performance on subsets of variables

Table 1 shows the results of the XGBoost algorithm on the 15 most important variables for males (M3) and 27 most important variables for females (F3). As expected from the SFFS procedure, the f1 scores for M3 and F3 were similar to M2 and F2. The f1 scores decreased by 1·8 percentage points for males and by 0·5 percentage points for females. Finally, we checked the performance of the algorithm on the nine most predictive common variables for both sexes (M4 and F4). The f1 scores were 72·9% for males and 72·4% for females, decreasing respectively by 2·6 and 3·7 percentage points compared to M2 and F2, and by 0·8 and 3·2 percentage points compared to M3 and F3. M4 and F4 were the models used for the calibration of the two scenarios.

### Scenarios

#### 1) 95% PLHIV know their status

Figures 4A and 4B show the PR-curves calculated on the test samples. For males, a sensitivity of 95% would require that 5,450 individuals out of 11,031 (49·4%) would need to be tested to identify 840 HIV positives out of the 883 PLHIV. The corresponding PPV is 15·4%; 7 individuals would therefore need to be tested to find one HIV positive person (number needed to test NNT). For females, 6,696 individuals out of 13,926 (48·1%) would need to be tested to find 1,522 HIV positives out of the 1,602 PLHIV. The PPV is 22·7% and the NNT is 5.

**Figure 4:**
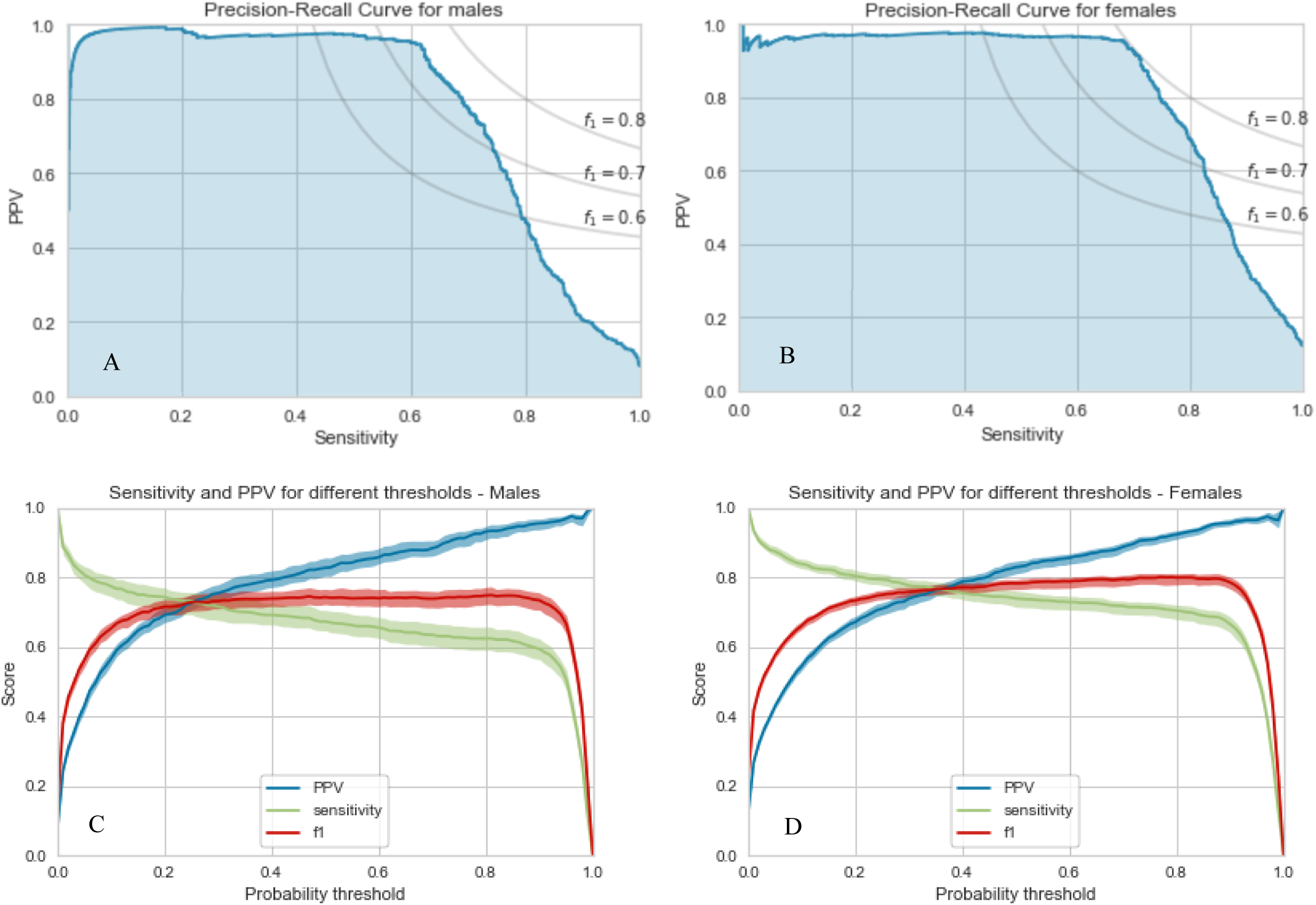
Precision-Recall Curves (A + B) and Threshold-Scores Curves (C+ D) for models with 9 variables (models M4 and F4) In addition, f1 iso-curves are shown for a typical range of f1 scores that we achieved with the models. Along these lines the f1 scores remain constant. Positive Predictive Value (PPV) Figure 4C and 4D: 95% Confidence Interval has been obtained using a bootstrap with n=50

#### 2) 95% or more probability of being HIV positive

Figures 4C and 4D show the TS-curves calculated on the test samples. Out of the 11,031 males and 13,926 females, 461 males (4·2%) and 862 females (6·2%) were identified as high-risk population. For males, 447 would have been correctly identified HIV positive out of the 883 PLHIV. For females, 833 would have been correctly identified HIV positive out of the 1,602 PLHIV.

## Discussion

Using large representative datasets with over 120,000 persons from ten East and Southern African countries, we were able to accurately predict the HIV status of individuals using demographic and socio-behavioural characteristics. Using all variables, XGBoost performed better than the three other algorithms on the test samples with a mean f1 score of 76·8% [95% CI 76·0%-77·6%] for males and 78.8% [78·2%-79·4%] for females. Our approach allowed us to select the nine most important predictor variables common for both sexes: geographic position (longitude, latitude and, altitude), current age, age of most recent partner, total lifetime number of sexual partners, years lived in current place of residence, condom used during last sexual intercourse with most recent partner and, wealth index. The performance of the algorithm using only these nine variables to predict HIV positivity was similar to that of the total dataset.

We also determined the direction of the association between predictor variables and HIV status. We confirmed a number of established HIV risk factors such as older age or older age of the most recent partner,^29^ a large number of sexual partners and, living in an urban area.^30^ Additionally, circumcision and breastfeeding were associated with a lower risk of HIV positivity. Unlike previous findings,^31^ condom use during the last sexual intercourse increased the probability of HIV positivity in our study. This seemingly counterintuitive finding may be the result of increased condom use in individuals who are already aware of their positive HIV status. The cross-sectional nature of our study limits our ability to investigate this further. We also identified risk factors for HIV infection which have rarely been investigated before. For example, an increased distance to water was positively associated with HIV infection in some persons, and negatively associated in others. This is in line with a previous study which showed that the risk of sexual assault of women, and hence the risk of HIV infection, increased when the time to reach a water source increased.^32^ However, longer time to get to water sources are more common in rural areas where HIV prevalence is generally lower, hence a decrease in risk of HIV positivity.

When applying these machine learning algorithms in real world settings, the trade-off between sensitivity (% of HIV positives identified) and PPV (yield) needs to be considered. A model with a sensitivity of 95% would be required to ensure that 95% of PLHIV know their status. In this first scenario, using a model with only nine predictors, we showed that the NNT was 5 (PPV of 15·4%) for males and 7 (PPV of 22·7%) for females. This represents approximately twice the PPV that would be achieved by general population testing. A previous systematic review of different testing strategies showed that NNTs ranged between 3 and 86 for community-based testing strategies and between 4 and 154 for facility-based testing strategies.^33^

In contrast, if targeted HIV case-finding strategies are implemented to increase the cost-effectiveness of testing strategies, a high PPV is important to ensure that the yield is high, and many of those tested are HIV positive. It is currently unknown if additional behavioural-based testing strategies can enhance or complement current targeted case-finding strategies such as index testing. Acceptable cut-offs for both sensitivity and PPV would need to be adapted for specific settings and for the desired testing coverage. For example, we defined a second scenario to simultaneously identify both high-risk HIV positive individuals for testing and high-risk HIV negative individuals for preventative services such as pre-exposure prophylaxis (PrEP).

To our knowledge, this study is the first to use machine learning methods to predict HIV in generalised HIV epidemic East and Southern African countries using routinely collected survey data. One of the limitations of this study is the generalizability of our findings. The distribution of risk factors varies between countries, and the accuracy of the prediction decreased for countries not used to train the algorithm. It is therefore not surprising, that geographic location of the residence (longitude, latitude and, altitude) were among the strongest predictors, since they were proxies for country-level differences. We were also limited by the available variables in our dataset, and as a result we were unable to consider differences in viral load suppression, health-care expenditure, specific HIV-related interventions and conflicts and wars. Additionally, although HIV testing was laboratory-based and not self-reported, some results were inconclusive and were discarded. A number of variables were self-reported and therefore subject to social desirability and recall bias. Missing values were imputed using multiple imputation, or directly within the extreme gradient boosting algorithm. However, the proportion of missing values was relatively small for most variables and both imputation methods gave similar results.

In conclusion, we were able to identify strong predictors of HIV positivity. Our findings may explain the spatial variability of HIV prevalence and can inform HIV testing strategies in resource-limited settings. While the implementation of a machine learning based risk score for targeted interventions was feasible in rural East Africa,^34^ the acceptability and use of potentially sensitive behavioural risk factors to directly identify individuals for HIV testing needs to be evaluated. Our algorithm performed well with only a limited number of variables, which do not require extensive interviews or questionnaires. This approach may be implemented by clinicians and community health care workers or utilised through additional HIV case-finding modalities such as call centres, social media and, self-testing initiatives. The availability of individual-level data on the association of various diseases with socio-behavioural characteristics is rapidly increasing. Advanced methods to analyse these large sources of data can help to prevent, diagnose and treat HIV and other diseases more efficiently.

## Data Availability

This is a study based on publicly available data from the Demographic and Health Surveys (DHS). The DHS Program is authorized to distribute, at no cost, unrestricted survey data files for legitimate academic research. Registration is required for access to data. https://dhsprogram.com/Data/

https://gitlab.com/Triphon/predicting_hiv_status

## Author’s contribution

EO, AM and, OK designed the study with support from SMM. EO wrote the code and performed the analysis with support from AM. EO, AM and, OK interpreted the results with support from JE and SMM. EO and RE reviewed the literature. EO, RE and, OK wrote the manuscript, which was reviewed by JE, SMM, and, AM.

## Acknowledgements

We acknowledge the support of the Swiss National Science Foundation (SNF professorship grant n° 163878 to O Keiser) which funded this study. We thank Antoine Flahault, Amaury Thiabaud, Danny Sheath and, Isotta Triulzi for helpful discussions and comments.

## Supplementary material

### Selection of variables

Datasets were resampled per country using sample weights from the HIV test results. We excluded individuals whose HIV status was “indeterminate” or “inconclusive” and individuals who reported that they never had sexual intercourse. We then removed variables with no variance and the ones containing more than 30% missing values. Finally, after additional encoding steps (e.g. creation of new aggregated variables or dummy coding of nominal variables), we manually removed an additional 77 non-informative variables for males and 122 for females (e.g. relating to metadata or information on how the survey was conducted), resulting in a final dataset of 55,151 males and 69,626 females with 84 and 122 variables, respectively. Overall 73 variables were common for both sexes (Table A2 and Table A3).

### Stratification, MICE imputation and Standardization (Figure 1 - step 1)

The stratification was done based on each sample HIV prevalence to ensure that the percentage of HIV positive individuals in the training and validation samples remained similar to the originals. We imputed missing values of each 80% training sample by multiple imputations using chained equations (MICE), and then applied the same imputation model to the corresponding test and left-out country samples. The regressions on the chained equations have been iterated ten times using the entire set of variables. The imputations have been performed five times and the results were averaged. Finally, the variables were standardized to a variance of one, ensuring that the penalization scheme is fair to all regressors.

**Table A1:** List of the Demographic and Health Surveys (DHS)

| Survey | Year | Country | Men |  | Women |  |
| --- | --- | --- | --- | --- | --- | --- |
|  |  |  | Individuals | Variables | Individuals | Variables |
| Standard DHS VII | 2015-2016 | Angola | 5,684 | 592 | 14,379 | 5,330 |
| Standard DHS VII | 2016-2017 | Burundi | 7,552 | 763 | 17,269 | 5,021 |
| Standard DHS VII | 2016 | Ethiopia | 12,688 | 590 | 15,683 | 5,695 |
| Standard DHS VII | 2014 | Lesotho | 2,931 | 627 | 6,621 | 3,748 |
| Standard DHS VII | 2015-2016 | Malawi | 7,478 | 571 | 24,562 | 4,934 |
| Standard AIS DHS VII | 2015 | Mozambique | 5,283 | 796 | 7,749 | 4,861 |
| Standard DHS VI | 2013 | Namibia | 4,481 | 641 | 10,018 | 4,180 |
| Standard DHS VII | 2014-2015 | Rwanda | 6,217 | 702 | 13,497 | 4,572 |
| Standard DHS VI | 2013-2014 | Zambia | 14,773 | 864 | 16,411 | 4,266 |
| Standard DHS VII | 2015 | Zimbabwe | 8,396 | 605 | 9,955 | 4,940 |
| Total |  |  | 75,483 |  | 136,144 |  |

**Table A2:** Pre-processing of variables.

| **Pre-process on the datasets** | **Men** |  | **Women** |  |
| --- | --- | --- | --- | --- |
|  | # of individuals | # of variables | # of individuals | # of variables |
| concatenation | 68,979 | 527 | 83,910 | 3,213 |
| 30% of missing values | 68,979 | 203 | 83,910 | 310 |
| no variance | 68,979 | 178 | 83,910 | 270 |
| duplicate variables | 68,979 | 173 | 83,910 | 261 |
| inconclusive HIV testing | 68,669 | 173 | 83,678 | 261 |
| no sexual intercourse | 55,151 | 173 | 69,626 | 261 |
| aggregation and removal | 55,151 | 84 | 69,626 | 122 |

**Table A3:**
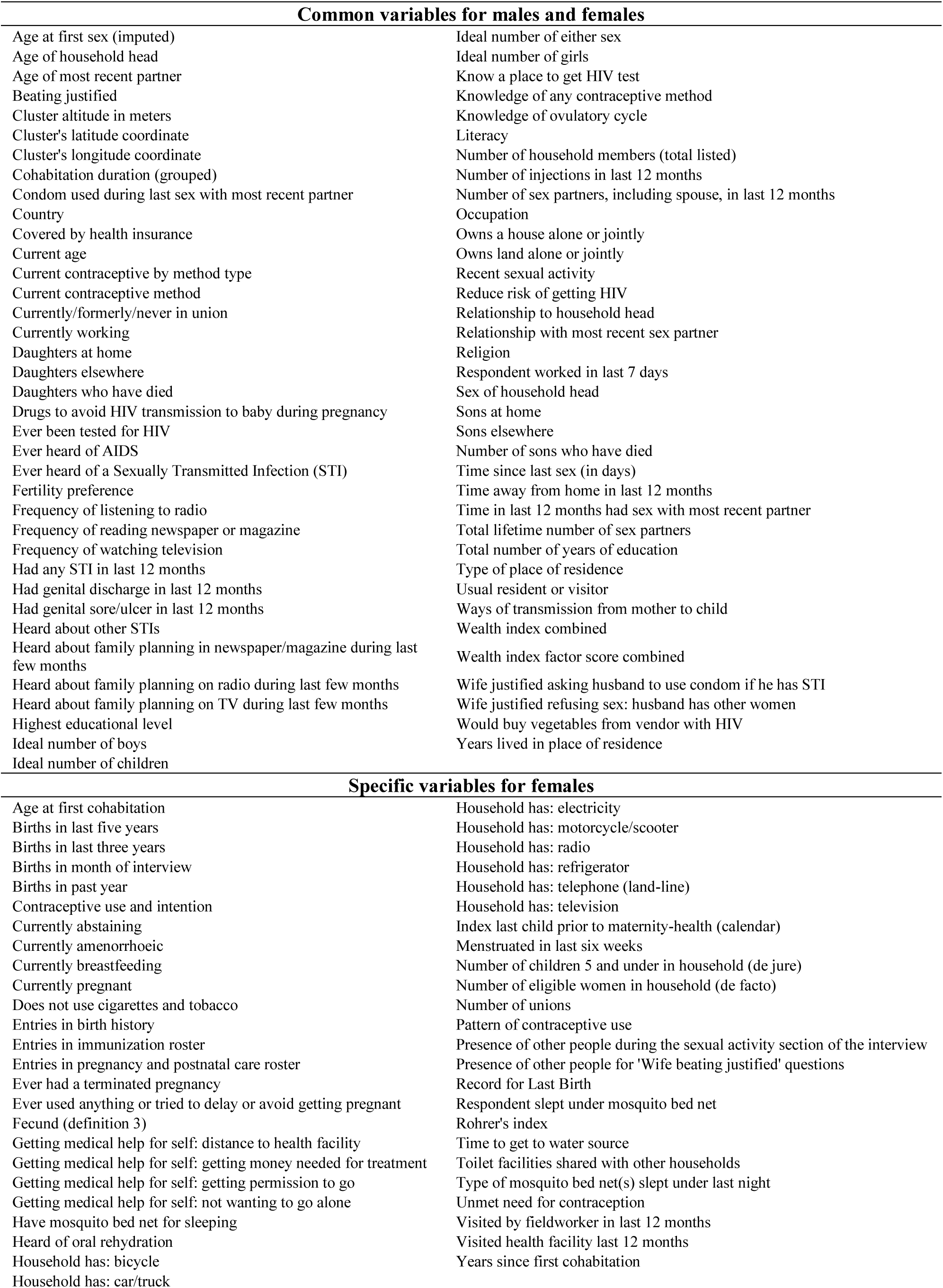
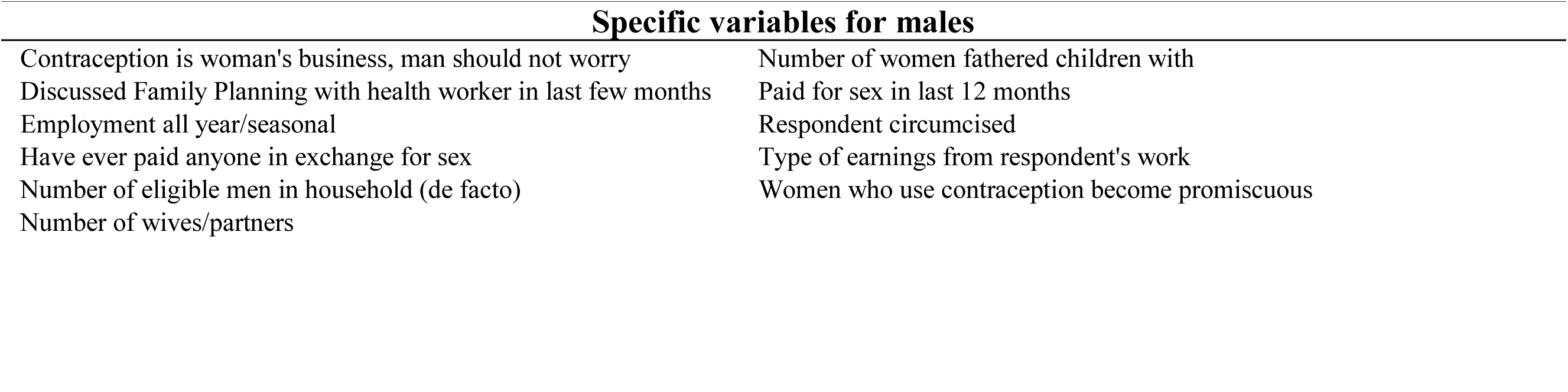
List of variables. Variable names correspond to the name in the Demographic and Health Survey (DHS).

| Common variables for males and females |  |
| --- | --- |
| Age at first sex (imputed) | Ideal number of either sex |
| Age of household head | Ideal number of girls |
| Age of most recent partner | Know a place to get HIV test |
| Beating justified | Knowledge of any contraceptive method |
| Cluster altitude in meters | Knowledge of ovulatory cycle |
| Cluster's latitude coordinate | Literacy |
| Cluster's longitude coordinate | Number of household members (total listed) |
| Cohabitation duration (grouped) | Number of injections in last 12 months |
| Condom used during last sex with most recent partner | Number of sex partners, including spouse, in last 12 months |
| Country | Occupation |
| Covered by health insurance | Owens a house alone or jointly |
| Current age | Owens land alone or jointly |
| Current contraceptive by method type | Recent sexual activity |
| Current contraceptive method | Reduce risk of getting HIV |
| Currently/formerly/never in union | Relationship to household head |
| Currently working | Relationship with most recent sex partner |
| Daughters at home | Religion |
| Daughters elsewhere | Respondent worked in last 7 days |
| Daughters who have died | Sex of household head |
| Drugs to avoid HIV transmission to baby during pregnancy | Sons at home |
| Ever been tested for HIV | Sons elsewhere |
| Ever heard of AIDS | Number of sons who have died |
| Ever heard of a Sexually Transmitted Infection (STI) | Time since last sex (in days) |
| Fertility preference | Time away from home in last 12 months |
| Frequency of listening to radio | Time in last 12 months had sex with most recent partner |
| Frequency of reading newspaper or magazine | Total lifetime number of sex partners |
| Frequency of watching television | Total number of years of education |
| Had any STI in last 12 months | Type of place of residence |
| Had genital discharge in last 12 months | Usual resident or visitor |
| Had genital sore/ulcer in last 12 months | Ways of transmission from mother to child |
| Heard about other STIs | Wealth index combined |
| Heard about family planning in newspaper/magazine during last few months | Wealth index factor score combined |
| Heard about family planning on radio during last few months | Wife justified asking husband to use condom if he has STI |
| Heard about family planning on TV during last few months | Wife justified refusing sex: husband has other women |
| Highest educational level | Would buy vegetables from vendor with HIV |
| Ideal number of boys | Years lived in place of residence |
| Ideal number of children |  |
| Specific variables for females |  |
| Age at first cohabitation | Household has: electricity |
| Births in last five years | Household has: motorcycle/scooter |
| Births in last three years | Household has: radio |
| Births in month of interview | Household has: refrigerator |
| Births in past year | Household has: telephone (land-line) |
| Contraceptive use and intention | Household has: television |
| Currently abstaining | Index last child prior to maternity-health (calendar) |
| Currently amenorrhoeic | Menstruated in last six weeks |
| Currently breastfeeding | Number of children 5 and under in household (de jure) |
| Currently pregnant | Number of eligible women in household (de facto) |
| Does not use cigarettes and tobacco | Number of unions |
| Entries in birth history | Pattern of contraceptive use |
| Entries in immunization roster | Presence of other people during the sexual activity section of the interview |
| Entries in pregnancy and postnatal care roster | Presence of other people for 'Wife beating justified' questions |
| Ever had a terminated pregnancy | Record for Last Birth |
| Ever used anything or tried to delay or avoid getting pregnant | Respondent slept under mosquito bed net |
| Fecund (definition 3) | Rohrer's index |
| Getting medical help for self: distance to health facility | Time to get to water source |
| Getting medical help for self: getting money needed for treatment | Toilet facilities shared with other households |
| Getting medical help for self: getting permission to go | Type of mosquito bed net(s) slept under last night |
| Getting medical help for self: not wanting to go alone | Unmet need for contraception |
| Have mosquito bed net for sleeping | Visited by fieldworker in last 12 months |
| Heard of oral rehydration | Visited health facility last 12 months |
| Household has: bicycle | Years since first cohabitation |
| Household has: car/truck |  |
| **Specific variables for males** |  |
| Contraception is woman's business, man should not worry | Number of women fathered children with |
| Discussed Family Planning with health worker in last few months | Paid for sex in last 12 months |
| Employment all year/seasonal | Respondent circumcised |
| Have ever paid anyone in exchange for sex | Type of earnings from respondent's work |
| Number of eligible men in household (de facto) | Women who use contraception become promiscuous |
| Number of wives/partners |  |

**Table A4:** Characteristics of Demographic and Health Survey (DHS) individuals.

|  |  | Male | Female |
| --- | --- | --- | --- |
| Total number of individuals |  | 55,151 | 69,626 |
| Current age, n (% of total) |  |  |  |
|  | 15-24 | 14,472 (26.2%) | 20,506 (29.5%) |
|  | 25-34 | 17,584 (31.9%) | 25,020 (35.9%) |
|  | 35-44 | 13,074 (23.7%) | 17,164 (24.7%) |
|  | 45-54 | 7,853 (14.2%) | 6,203 (8.9%) |
|  | 55-64 | 2,168 (3.9%) | 733 (1.1%) |
| Country of origin, n (% of total) |  |  |  |
|  | Angola | 4,611 (8.4%) | 6,020 (8.6%) |
|  | Burundi | 5,180 (9.4%) | 6,007 (8.6%) |
|  | Ethiopia | 7,955 (14.4%) | 11,162 (16.0%) |
|  | Lesotho | 2,438 (4.4%) | 2,917 (4.2%) |
|  | Malawi | 5,657 (10.3%) | 6,895 (9.9%) |
|  | Mozambique | 4,053 (7.3%) | 6,472 (9.3%) |
|  | Namibia | 3,326 (6.0%) | 4,431 (6.4%) |
|  | Rwanda | 4,613 (8.4%) | 4,948 (7.1%) |
|  | Zambia | 11,617 (21.1%) | 13,453 (19.3%) |
|  | Zimbabwe | 5,701 (10.3%) | 7,321 (10.5%) |
| HIV positive by country, n (% of individuals within country) |  |  |  |
|  | Angola | 48 (1.0%) | 164 (2.7%) |
|  | Burundi | 49 (0.9%) | 92 (1.5%) |
|  | Ethiopia | 66 (0.8%) | 172 (1.5%) |
|  | Lesotho | 531 (21.8%) | 970 (33.3%) |
|  | Malawi | 445 (7.9%) | 837 (12.1%) |
|  | Mozambique | 434 (10.7%) | 1,002 (15.5%) |
|  | Namibia | 433 (13.0%) | 810 (18.3%) |
|  | Rwanda | 157 (3.4%) | 264 (5.3%) |
|  | Zambia | 1,494 (12.9%) | 2,235 (16.6%) |
|  | Zimbabwe | 760 (13.3%) | 1,465 (20.0%) |
| Type of residence, n (% of total) |  |  |  |
|  | Urban | 19,196 (34.8%) | 23,501 (33.8%) |
|  | Rural | 35,955 (65.2%) | 46,125 (66.2%) |

**Table A5i:** Results of the XGBoost algorithm per sex for the validation, test and, left-out samples.

| Country | Metric | Males |  |  |  | Females |  |  |  |
| --- | --- | --- | --- | --- | --- | --- | --- | --- | --- |
|  |  | f1 score | Sensitivity | PPV | Prevalence | f1 score | Sensitivity | PPV | Prevalence |
| Angola | Validation | 73.3% ($\pm 2.3\%$) | 70.7% ($\pm 3.7\%$) | 76.2% ($\pm 2.4\%$) | 8.6% | 74.9% ($\pm 0.9\%$) | 72.6% ($\pm 1.6\%$) | 77.4% ($\pm 0.5\%$) | 12.3% |
|  | Test | 75.9% | 72.2% | 80.1% | 8.6% | 78.9% | 76.1% | 81.9% | 12.3% |
|  | Left-out | 6.6% | 16.7% | 4.1% | 1.0% | 12.2% | 12.8% | 11.6% | 2.7% |
| Burundi | Validation | 73.8% ($\pm 2.0\%$) | 70.7% ($\pm 2.3\%$) | 77.3% ($\pm 3.6\%$) | 8.7% | 74.5% ($\pm 1.0\%$) | 74.5% ($\pm 1.6\%$) | 74.6% ($\pm 1.2\%$) | 12.4% |
|  | Test | 75.6% | 73.0% | 78.4% | 8.7% | 79.0% | 78.1% | 79.9% | 12.4% |
|  | Left-out | 17.1% | 14.3% | 21.2% | 0.9% | 17.6% | 22.8% | 14.3% | 1.5% |
| Ethiopia | Validation | 72.7% ($\pm 2.4\%$) | 66.3% ($\pm 4.0\%$) | 80.6% ($\pm 1.3\%$) | 9.2% | 74.9% ($\pm 0.6\%$) | 71.8% ($\pm 1.0\%$) | 78.3% ($\pm 1.1\%$) | 13.4% |
|  | Test | 78.9% | 74.8% | 83.4% | 9.2% | 80.0% | 78.1% | 81.9% | 13.4% |
|  | Left-out | 12.5% | 7.6% | 35.7% | 0.8% | 5.3% | 3.5% | 11.1% | 1.5% |
| Lesotho | Validation | 72.9% ($\pm 1.7\%$) | 68.4% ($\pm 3.6\%$) | 78.1% ($\pm 2.3\%$) | 7.4% | 74.7% ($\pm 1.1\%$) | 71.0% ($\pm 2.7\%$) | 78.9% ($\pm 1.5\%$) | 10.6% |
|  | Test | 76.4% | 71.9% | 81.4% | 7.4% | 78.0% | 74.8% | 81.6% | 10.6% |
|  | Left-out | 32.8% | 22.6% | 60.0% | 21.8% | 47.5% | 37.5% | 64.8% | 33.3% |
| Malawi | Validation | 73.4% ($\pm 2.7\%$) | 69.5% ($\pm 2.3\%$) | 77.7% ($\pm 3.8\%$) | 8.0% | 75.9% ($\pm 1.5\%$) | 73.2% ($\pm 1.4\%$) | 78.9% ($\pm 1.8\%$) | 11.4% |
|  | Test | 77.3% | 73.4% | 81.7% | 8.0% | 78.6% | 76.1% | 81.3% | 11.4% |
|  | Left-out | 24.9% | 18.9% | 36.7% | 7.9% | 32.8% | 30.1% | 36.1% | 12.1% |
| Mozambique | Validation | 74.1% ($\pm 1.7\%$) | 68.3% ($\pm 2.6\%$) | 81.0% ($\pm 3.3\%$) | 7.8% | 76.6% ($\pm 1.0\%$) | 75.4% ($\pm 1.1\%$) | 78.0% ($\pm 1.3\%$) | 11.1% |
|  | Test | 75.5% | 68.4% | 84.4% | 7.8% | 80.5% | 79.2% | 81.9% | 11.1% |
|  | Left-out | 18.9% | 12.9% | 35.4% | 10.7% | 31.2% | 25.8% | 39.2% | 15.5% |
| Namibia | Validation | 73.6% ($\pm 1.7\%$) | 69.6% ($\pm 1.2\%$) | 78.1% ($\pm 3.3\%$) | 7.7% | 75.4% ($\pm 0.9\%$) | 73.4% ($\pm 0.9\%$) | 77.5% ($\pm 1.4\%$) | 11.0% |
|  | Test | 77.4% | 73.8% | 81.3% | 7.7% | 78.2% | 77.8% | 78.6% | 11.0% |
|  | Left-out | 31.2% | 27.9% | 35.3% | 13.0% | 41.8% | 41.5% | 42.1% | 18.3% |
| Rwanda | Validation | 72.9% ($\pm 2.5\%$) | 69.7% ($\pm 2.7\%$) | 76.4% ($\pm 2.6\%$) | 8.4% | 75.5% ($\pm 1.2\%$) | 74.7% ($\pm 1.6\%$) | 76.3% ($\pm 1.1\%$) | 12.0% |
|  | Test | 78.0% | 75.6% | 80.6% | 8.4% | 79.3% | 79.7% | 78.9% | 12.0% |
|  | Left-out | 11.3% | 6.4% | 50.0% | 3.4% | 20.8% | 16.7% | 27.5% | 5.3% |
| Zambia | Validation | 72.8% ($\pm 2.3\%$) | 69.4% ($\pm 2.0\%$) | 76.6% ($\pm 3.5\%$) | 6.7% | 74.2% ($\pm 2.1\%$) | 73.7% ($\pm 1.9\%$) | 74.8% ($\pm 2.6\%$) | 10.3% |
|  | Test | 76.7% | 73.0% | 80.7% | 6.7% | 77.7% | 77.0% | 78.5% | 10.3% |
|  | Left-out | 22.3% | 14.1% | 53.0% | 12.9% | 41.5% | 37.1% | 47.0% | 16.6% |
| Zimbabwe | Validation | 73.2% ($\pm 1.4\%$) | 66.9% ($\pm 1.6\%$) | 80.7% ($\pm 1.6\%$) | 7.4% | 75.0% ($\pm 0.7\%$) | 73.1% ($\pm 1.6\%$) | 77.1% ($\pm 1.1\%$) | 10.5% |
|  | Test | 76.3% | 70.7% | 82.9% | 7.4% | 78.2% | 77.4% | 79.0% | 10.5% |
|  | Left-out | 31.3% | 27.2% | 36.9% | 13.3% | 47.0% | 41.3% | 54.5% | 20.0% |

Positive Predictive Value (PPV)
( $\pm \%$ ): 95% Confidence Interval

**Table A5ii:** Results of the Support Vector Machine (SVM) algorithm per sex for the validation, test and, left-out samples.

| Country | Metric | Males |  |  |  | Females |  |  |  |
| --- | --- | --- | --- | --- | --- | --- | --- | --- | --- |
|  |  | f1 score | Sensitivity | PPV | Prevalence | f1 score | Sensitivity | PPV | Prevalence |
| Angola | Validation | 64.1% ($\pm 2.2\%$) | 66.4% ($\pm 2.0\%$) | 62.0% ($\pm 2.9\%$) | 8.6% | 70.3% ($\pm 1.2\%$) | 69.3% ($\pm 1.1\%$) | 71.4% ($\pm 1.8\%$) | 12.3% |
|  | Test | 70.1% | 71.6% | 68.7% | 8.6% | 74.3% | 73.3% | 75.3% | 12.3% |
|  | Left-out | 10.2% | 22.9% | 6.5% | 1.0% | 4.9% | 7.9% | 3.5% | 2.7% |
| Burundi | Validation | 64.9% ($\pm 0.6\%$) | 66.4% ($\pm 1.6\%$) | 63.6% ($\pm 2.6\%$) | 8.7% | 71.0% ($\pm 1.2\%$) | 68.5% ($\pm 1.2\%$) | 73.7% ($\pm 2.8\%$) | 12.4% |
|  | Test | 68.5% | 69.5% | 67.5% | 8.7% | 75.3% | 72.1% | 78.9% | 12.4% |
|  | Left-out | 11.2% | 22.4% | 7.4% | 0.9% | 8.0% | 12.0% | 6.0% | 1.5% |
| Ethiopia | Validation | 63.8% ($\pm 2.7\%$) | 64.5% ($\pm 3.9\%$) | 63.1% ($\pm 2.4\%$) | 9.2% | 70.1% ($\pm 1.2\%$) | 67.7% ($\pm 1.1\%$) | 72.8% ($\pm 1.8\%$) | 13.4% |
|  | Test | 70.4% | 73.2% | 67.8% | 9.2% | 75.1% | 74.4% | 75.8% | 13.4% |
|  | Left-out | 3.2% | 7.6% | 2.0% | 0.8% | 18.9% | 23.8% | 15.6% | 1.5% |
| Lesotho | Validation | 64.5% ($\pm 0.9\%$) | 65.6% ($\pm 2.2\%$) | 63.5% ($\pm 2.0\%$) | 7.4% | 69.7% ($\pm 1.3\%$) | 67.4% ($\pm 1.6\%$) | 72.1% ($\pm 1.9\%$) | 10.6% |
|  | Test | 69.4% | 70.5% | 68.3% | 7.4% | 73.3% | 71.2% | 75.4% | 10.6% |
|  | Left-out | 22.3% | 15.4% | 39.8% | 21.8% | 43.9% | 37.3% | 53.3% | 33.3% |
| Malawi | Validation | 65.2% ($\pm 2.3\%$) | 65.5% ($\pm 3.3\%$) | 65.0% ($\pm 1.5\%$) | 8.0% | 70.8% ($\pm 1.2\%$) | 68.8% ($\pm 1.5\%$) | 72.9% ($\pm 1.2\%$) | 11.4% |
|  | Test | 69.9% | 70.5% | 69.2% | 8.0% | 75.0% | 73.0% | 77.1% | 11.4% |
|  | Left-out | 21.7% | 17.8% | 27.8% | 7.9% | 25.9% | 21.7% | 32.0% | 12.1% |
| Mozambique | Validation | 66.6% ($\pm 1.2\%$) | 67.7% ($\pm 1.7\%$) | 65.6% ($\pm 1.5\%$) | 7.8% | 72.4% ($\pm 1.9\%$) | 70.2% ($\pm 2.1\%$) | 74.7% ($\pm 2.3\%$) | 11.1% |
|  | Test | 68.7% | 69.8% | 67.7% | 7.8% | 77.1% | 74.2% | 80.2% | 11.1% |
|  | Left-out | 11.4% | 8.5% | 17.4% | 10.7% | 24.9% | 21.6% | 29.5% | 15.5% |
| Namibia | Validation | 66.4% ($\pm 1.5\%$) | 66.8% ($\pm 3.1\%$) | 66.0% ($\pm 0.6\%$) | 7.7% | 70.1% ($\pm 1.9\%$) | 67.5% ($\pm 2.1\%$) | 72.8% ($\pm 2.1\%$) | 11.0% |
|  | Test | 69.8% | 70.1% | 69.5% | 7.7% | 74.1% | 71.7% | 76.7% | 11.0% |
|  | Left-out | 15.5% | 11.1% | 25.9% | 13.0% | 29.6% | 27.8% | 31.7% | 18.3% |
| Rwanda | Validation | 65.2% ($\pm 2.4\%$) | 65.9% ($\pm 3.0\%$) | 64.6% ($\pm 2.7\%$) | 8.4% | 70.8% ($\pm 1.2\%$) | 68.7% ($\pm 1.4\%$) | 73.1% ($\pm 2.0\%$) | 12.0% |
|  | Test | 71.1% | 73.6% | 68.8% | 8.4% | 75.0% | 73.7% | 76.4% | 12.0% |
|  | Left-out | 19.5% | 22.9% | 17.0% | 3.4% | 15.8% | 13.6% | 18.8% | 5.3% |
| Zambia | Validation | 65.0% ($\pm 2.2\%$) | 64.8% ($\pm 2.0\%$) | 65.2% ($\pm 2.4\%$) | 6.7% | 70.2% ($\pm 1.5\%$) | 67.1% ($\pm 1.6\%$) | 73.7% ($\pm 2.2\%$) | 10.3% |
|  | Test | 66.9% | 67.2% | 66.6% | 6.7% | 74.0% | 71.0% | 77.4% | 10.3% |
|  | Left-out | 21.1% | 15.5% | 33.2% | 12.9% | 31.3% | 23.5% | 47.0% | 16.6% |
| Zimbabwe | Validation | 63.7% ($\pm 1.8\%$) | 64.5% ($\pm 1.9\%$) | 62.8% ($\pm 1.9\%$) | 7.4% | 70.4% ($\pm 1.0\%$) | 67.8% ($\pm 2.1\%$) | 73.2% ($\pm 2.1\%$) | 10.5% |
|  | Test | 67.3% | 67.6% | 66.9% | 7.4% | 72.6% | 70.1% | 75.3% | 10.5% |
|  | Left-out | 18.3% | 14.6% | 24.4% | 13.3% | 19.7% | 12.5% | 46.8% | 20.0% |

Positive Predictive Value (PPV)
( $\pm \%$ ): 95% Confidence Interval

**Table A5iii:** Results of the Elastic Net algorithm per sex for the validation, test and, left-out samples.

| Country | Metric | Males |  |  |  | Females |  |  |  |
| --- | --- | --- | --- | --- | --- | --- | --- | --- | --- |
|  |  | f1 score | Sensitivity | PPV | Prevalence | f1 score | Sensitivity | PPV | Prevalence |
| Angola | Validation | 33.3% ($\pm 1.6\%$) | 76.8% ($\pm 3.4\%$) | 21.2% ($\pm 1.1\%$) | 8.6% | 42.2% ($\pm 0.7\%$) | 77.0% ($\pm 1.6\%$) | 29.0% ($\pm 0.5\%$) | 12.3% |
|  | Test | 33.7% | 77.6% | 21.5% | 8.6% | 43.1% | 77.8% | 29.8% | 12.3% |
|  | Left-out | 3.5% | 56.2% | 1.8% | 1.0% | 10.1% | 65.2% | 5.5% | 2.7% |
| Burundi | Validation | 33.7% ($\pm 1.0\%$) | 76.8% ($\pm 3.3\%$) | 21.6% ($\pm 0.6\%$) | 8.7% | 42.1% ($\pm 1.8\%$) | 76.4% ($\pm 2.7\%$) | 29.1% ($\pm 1.4\%$) | 12.4% |
|  | Test | 33.1% | 77.1% | 21.1% | 8.7% | 41.8% | 76.2% | 28.8% | 12.4% |
|  | Left-out | 6.8% | 63.3% | 3.6% | 0.9% | 13.1% | 75.0% | 7.2% | 1.5% |
| Ethiopia | Validation | 34.4% ($\pm 0.7\%$) | 75.3% ($\pm 1.6\%$) | 22.3% ($\pm 0.6\%$) | 9.2% | 43.2% ($\pm 1.3\%$) | 75.0% ($\pm 2.4\%$) | 30.3% ($\pm 1.0\%$) | 13.4% |
|  | Test | 33.3% | 72.2% | 21.6% | 9.2% | 43.6% | 75.9% | 30.6% | 13.4% |
|  | Left-out | 4.7% | 47.0% | 2.5% | 0.8% | 19.2% | 57.6% | 11.5% | 1.5% |
| Lesotho | Validation | 30.3% ($\pm 0.9\%$) | 78.1% ($\pm 2.2\%$) | 18.8% ($\pm 0.5\%$) | 7.4% | 38.8% ($\pm 0.8\%$) | 77.7% ($\pm 1.7\%$) | 25.9% ($\pm 0.6\%$) | 10.6% |
|  | Test | 29.8% | 77.3% | 18.5% | 7.4% | 38.6% | 78.1% | 25.6% | 10.6% |
|  | Left-out | 37.0% | 30.7% | 46.7% | 21.8% | 60.7% | 85.7% | 46.9% | 33.3% |
| Malawi | Validation | 32.9% ($\pm 0.6\%$) | 79.1% ($\pm 1.1\%$) | 20.8% ($\pm 0.5\%$) | 8.0% | 41.9% ($\pm 1.6\%$) | 78.9% ($\pm 2.6\%$) | 28.5% ($\pm 1.1\%$) | 11.4% |
|  | Test | 33.2% | 80.5% | 20.9% | 8.0% | 41.9% | 79.0% | 28.5% | 11.4% |
|  | Left-out | 31.1% | 49.9% | 22.6% | 7.9% | 37.8% | 58.9% | 27.8% | 12.1% |
| Mozambique | Validation | 32.4% ($\pm 0.4\%$) | 78.5% ($\pm 1.3\%$) | 20.4% ($\pm 0.2\%$) | 7.8% | 42.2% ($\pm 0.6\%$) | 79.1% ($\pm 1.6\%$) | 28.8% ($\pm 0.5\%$) | 11.1% |
|  | Test | 33.9% | 78.7% | 21.6% | 7.8% | 42.2% | 79.7% | 28.7% | 11.1% |
|  | Left-out | 23.6% | 29.0% | 19.9% | 10.7% | 30.1% | 80.5% | 18.5% | 15.5% |
| Namibia | Validation | 32.2% ($\pm 0.6\%$) | 78.4% ($\pm 1.4\%$) | 20.3% ($\pm 0.5\%$) | 7.7% | 41.6% ($\pm 1.3\%$) | 79.0% ($\pm 1.0\%$) | 28.2% ($\pm 1.1\%$) | 11.0% |
|  | Test | 32.3% | 76.9% | 20.4% | 7.7% | 41.0% | 78.0% | 27.8% | 11.0% |
|  | Left-out | 32.2% | 59.8% | 22.0% | 13.0% | 37.2% | 83.1% | 24.0% | 18.3% |
| Rwanda | Validation | 32.9% ($\pm 0.6\%$) | 78.3% ($\pm 2.0\%$) | 20.8% ($\pm 0.4\%$) | 8.4% | 41.9% ($\pm 1.0\%$) | 78.1% ($\pm 1.1\%$) | 28.6% ($\pm 0.9\%$) | 12.0% |
|  | Test | 32.7% | 78.5% | 20.7% | 8.4% | 43.0% | 80.1% | 29.4% | 12.0% |
|  | Left-out | 15.3% | 84.7% | 8.4% | 3.4% | 26.2% | 54.9% | 17.2% | 5.3% |
| Zambia | Validation | 31.7% ($\pm 1.5\%$) | 80.5% ($\pm 1.3\%$) | 19.8% ($\pm 1.1\%$) | 6.7% | 40.5% ($\pm 0.9\%$) | 80.0% ($\pm 1.1\%$) | 27.1% ($\pm 0.7\%$) | 10.3% |
|  | Test | 31.7% | 79.7% | 19.8% | 6.7% | 40.3% | 79.1% | 27.0% | 10.3% |
|  | Left-out | 32.5% | 70.5% | 21.2% | 12.9% | 43.8% | 72.8% | 31.4% | 16.6% |
| Zimbabwe | Validation | 31.1% ($\pm 0.6\%$) | 78.2% ($\pm 1.5\%$) | 19.4% ($\pm 0.4\%$) | 7.4% | 39.3% ($\pm 1.1\%$) | 78.1% ($\pm 1.7\%$) | 26.3% ($\pm 1.0\%$) | 10.5% |
|  | Test | 32.2% | 80.3% | 20.2% | 7.4% | 39.4% | 77.8% | 26.4% | 10.5% |
|  | Left-out | 27.1% | 63.4% | 17.2% | 13.3% | 47.8% | 68.2% | 36.8% | 20.0% |

Positive Predictive Value (PPV)
( $\pm \%$ ): 95% Confidence Interval

**Table A5iv:**
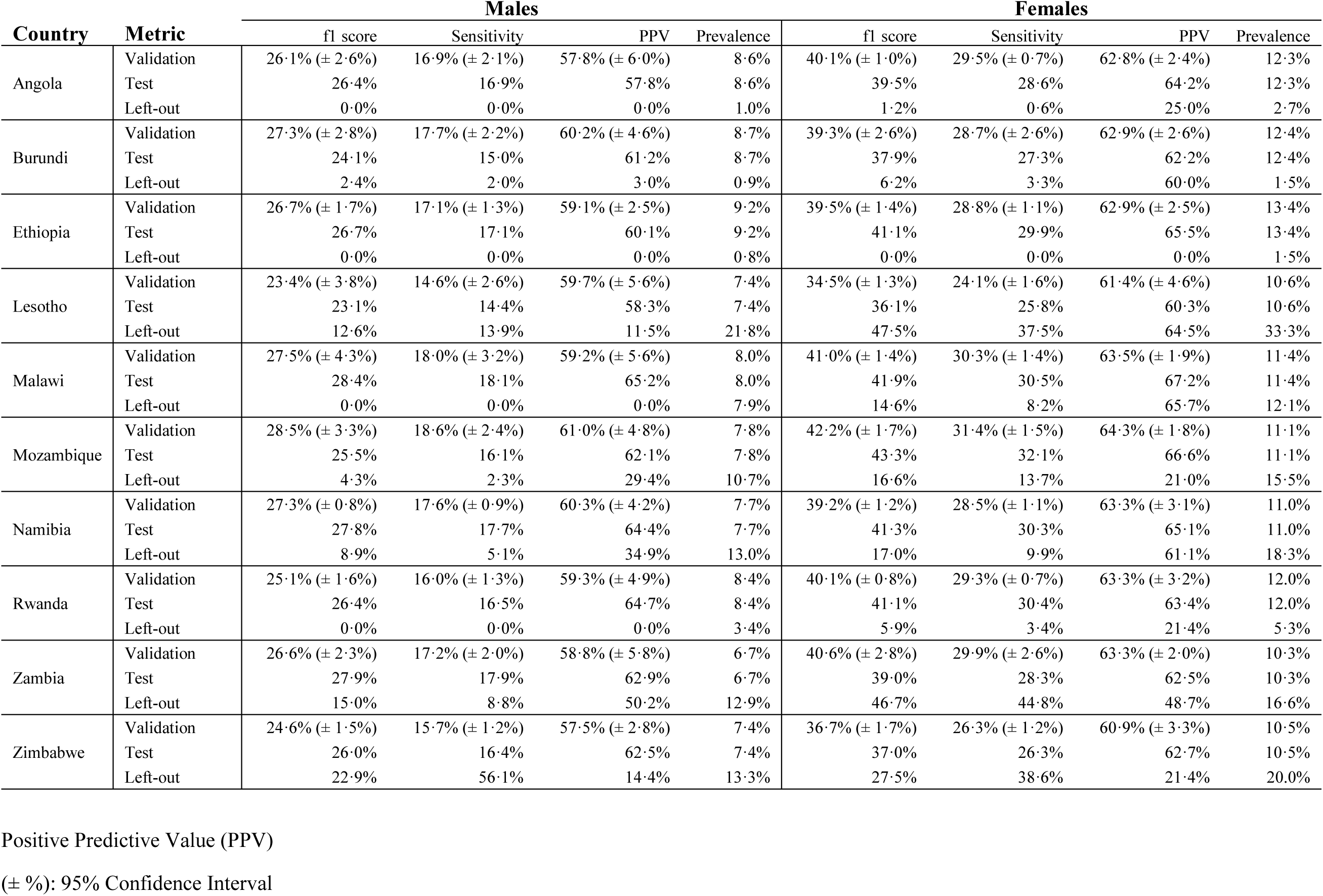
Results of the Generalized Additive Model (GAM) algorithm per sex for the validation, test and, left-out samples.

| Country | Metric | Males |  |  |  | Females |  |  |  |
| --- | --- | --- | --- | --- | --- | --- | --- | --- | --- |
|  |  | f1 score | Sensitivity | PPV | Prevalence | f1 score | Sensitivity | PPV | Prevalence |
| Angola | Validation | 26.1% ($\pm 2.6\%$) | 16.9% ($\pm 2.1\%$) | 57.8% ($\pm 6.0\%$) | 8.6% | 40.1% ($\pm 1.0\%$) | 29.5% ($\pm 0.7\%$) | 62.8% ($\pm 2.4\%$) | 12.3% |
|  | Test | 26.4% | 16.9% | 57.8% | 8.6% | 39.5% | 28.6% | 64.2% | 12.3% |
|  | Left-out | 0.0% | 0.0% | 0.0% | 1.0% | 1.2% | 0.6% | 25.0% | 2.7% |
| Burundi | Validation | 27.3% ($\pm 2.8\%$) | 17.7% ($\pm 2.2\%$) | 60.2% ($\pm 4.6\%$) | 8.7% | 39.3% ($\pm 2.6\%$) | 28.7% ($\pm 2.6\%$) | 62.9% ($\pm 2.6\%$) | 12.4% |
|  | Test | 24.1% | 15.0% | 61.2% | 8.7% | 37.9% | 27.3% | 62.2% | 12.4% |
|  | Left-out | 2.4% | 2.0% | 3.0% | 0.9% | 6.2% | 3.3% | 60.0% | 1.5% |
| Ethiopia | Validation | 26.7% ($\pm 1.7\%$) | 17.1% ($\pm 1.3\%$) | 59.1% ($\pm 2.5\%$) | 9.2% | 39.5% ($\pm 1.4\%$) | 28.8% ($\pm 1.1\%$) | 62.9% ($\pm 2.5\%$) | 13.4% |
|  | Test | 26.7% | 17.1% | 60.1% | 9.2% | 41.1% | 29.9% | 65.5% | 13.4% |
|  | Left-out | 0.0% | 0.0% | 0.0% | 0.8% | 0.0% | 0.0% | 0.0% | 1.5% |
| Lesotho | Validation | 23.4% ($\pm 3.8\%$) | 14.6% ($\pm 2.6\%$) | 59.7% ($\pm 5.6\%$) | 7.4% | 34.5% ($\pm 1.3\%$) | 24.1% ($\pm 1.6\%$) | 61.4% ($\pm 4.6\%$) | 10.6% |
|  | Test | 23.1% | 14.4% | 58.3% | 7.4% | 36.1% | 25.8% | 60.3% | 10.6% |
|  | Left-out | 12.6% | 13.9% | 11.5% | 21.8% | 47.5% | 37.5% | 64.5% | 33.3% |
| Malawi | Validation | 27.5% ($\pm 4.3\%$) | 18.0% ($\pm 3.2\%$) | 59.2% ($\pm 5.6\%$) | 8.0% | 41.0% ($\pm 1.4\%$) | 30.3% ($\pm 1.4\%$) | 63.5% ($\pm 1.9\%$) | 11.4% |
|  | Test | 28.4% | 18.1% | 65.2% | 8.0% | 41.9% | 30.5% | 67.2% | 11.4% |
|  | Left-out | 0.0% | 0.0% | 0.0% | 7.9% | 14.6% | 8.2% | 65.7% | 12.1% |
| Mozambique | Validation | 28.5% ($\pm 3.3\%$) | 18.6% ($\pm 2.4\%$) | 61.0% ($\pm 4.8\%$) | 7.8% | 42.2% ($\pm 1.7\%$) | 31.4% ($\pm 1.5\%$) | 64.3% ($\pm 1.8\%$) | 11.1% |
|  | Test | 25.5% | 16.1% | 62.1% | 7.8% | 43.3% | 32.1% | 66.6% | 11.1% |
|  | Left-out | 4.3% | 2.3% | 29.4% | 10.7% | 16.6% | 13.7% | 21.0% | 15.5% |
| Namibia | Validation | 27.3% ($\pm 0.8\%$) | 17.6% ($\pm 0.9\%$) | 60.3% ($\pm 4.2\%$) | 7.7% | 39.2% ($\pm 1.2\%$) | 28.5% ($\pm 1.1\%$) | 63.3% ($\pm 3.1\%$) | 11.0% |
|  | Test | 27.8% | 17.7% | 64.4% | 7.7% | 41.3% | 30.3% | 65.1% | 11.0% |
|  | Left-out | 8.9% | 5.1% | 34.9% | 13.0% | 17.0% | 9.9% | 61.1% | 18.3% |
| Rwanda | Validation | 25.1% ($\pm 1.6\%$) | 16.0% ($\pm 1.3\%$) | 59.3% ($\pm 4.9\%$) | 8.4% | 40.1% ($\pm 0.8\%$) | 29.3% ($\pm 0.7\%$) | 63.3% ($\pm 3.2\%$) | 12.0% |
|  | Test | 26.4% | 16.5% | 64.7% | 8.4% | 41.1% | 30.4% | 63.4% | 12.0% |
|  | Left-out | 0.0% | 0.0% | 0.0% | 3.4% | 5.9% | 3.4% | 21.4% | 5.3% |
| Zambia | Validation | 26.6% ($\pm 2.3\%$) | 17.2% ($\pm 2.0\%$) | 58.8% ($\pm 5.8\%$) | 6.7% | 40.6% ($\pm 2.8\%$) | 29.9% ($\pm 2.6\%$) | 63.3% ($\pm 2.0\%$) | 10.3% |
|  | Test | 27.9% | 17.9% | 62.9% | 6.7% | 39.0% | 28.3% | 62.5% | 10.3% |
|  | Left-out | 15.0% | 8.8% | 50.2% | 12.9% | 46.7% | 44.8% | 48.7% | 16.6% |
| Zimbabwe | Validation | 24.6% ($\pm 1.5\%$) | 15.7% ($\pm 1.2\%$) | 57.5% ($\pm 2.8\%$) | 7.4% | 36.7% ($\pm 1.7\%$) | 26.3% ($\pm 1.2\%$) | 60.9% ($\pm 3.3\%$) | 10.5% |
|  | Test | 26.0% | 16.4% | 62.5% | 7.4% | 37.0% | 26.3% | 62.7% | 10.5% |
|  | Left-out | 22.9% | 56.1% | 14.4% | 13.3% | 27.5% | 38.6% | 21.4% | 20.0% |

Positive Predictive Value (PPV)
( $\pm \%$ ): 95% Confidence Interval

## Python libraries

- Matplotlib 3.1.1
- Mlxtend 0.17.0
- Numpy 1.16.5
- Pandas 0.25.1
- Pathlib 1.0.1
- Pyshp 2.1.0
- Pygam 0.8.0
- Scikit-learn 0.21.3
- Scipy 1.3.1
- Seaborn 0.9.0
- Shap 0.30.1
- Xgboost 0.90
- Yellowbrick 1.0.1

Some of the computations were done on the Baobab cluster of the University of Geneva.

